# Modeling quantitative traits for COVID-19 case reports

**DOI:** 10.1101/2020.06.18.20135103

**Authors:** Núria Queralt-Rosinach, Susan M. Bello, Robert Hoehndorf, Claus Weiland, Philippe Rocca-Serra, Paul N. Schofield

## Abstract

Medical practitioners record the condition status of a patient through qualitative and quantitative observations. The measurement of vital signs and molecular parameters in the clinics gives a complementary description of abnormal phenotypes associated with the progression of a disease. The Clinical Measurement Ontology (CMO) is used to standardize annotations of these measurable traits. However, researchers have no way to describe how these quantitative traits relate to phenotype concepts in a machine-readable manner. Using the WHO clinical case report form standard for the COVID-19 pandemic, we modeled quantitative traits and developed OWL axioms to formally relate clinical measurement terms with anatomical, biomolecular entities and phenotypes annotated with the Uber-anatomy ontology (Uberon), Chemical Entities of Biological Interest (ChEBI) and the Phenotype and Trait Ontology (PATO) biomedical ontologies. The formal description of these relations allows interoperability between clinical and biological descriptions, and facilitates automated reasoning for analysis of patterns over quantitative and qualitative biomedical observations.

## Introduction

The worldwide COVID-19 pandemic has brought into focus the need in research to have patient data more available and accessible to get new insights more efficiently and rapidly. Clinicians monitor biomolecular concentrations, other physiological signs, and symptoms manifested in different organ systems of the patient at different points in time. These clinical measures are very valuable data because they give intrinsic information about the underlying biological mechanism and patient disease trajectory that could be used to make informed and tailored therapeutic decisions. One problem that clinicians face is how to interpret patient laboratory results and quantitative traits, such as serum electrolyte concentrations or blood pressure, and link to their significance at a phenotypic conceptual level. To bridge this gap, computational biologists use ontologies to encode knowledge about how altered gene products and molecular processes affect cellular and tissue functions that ultimately are expressed as abnormal phenotypes and diseases. The life science community has been developing different ontologies to represent both molecular biology, clinical measures, and disease phenotypes. For instance the CMO [1] is designed to encode morphological and physiological measurement data generated from clinical and model organism research and health records. However, there is still the need to link this quantitative measurement standard to phenotypic ontologies. It is necessary to establish if a measurement falls into an abnormal range, if so what kind of phenotype might result, and what the consequences of the phenotype might be, either diagnostically or prognostically. Assignment of some phenotypes requires several measurements to be abnormal; these then need to be integrated. A clinician can then understand that given this phenotype, possibly along with others, a diagnosis may be made of a specific disease which then carries with it diagnostic and prognostic knowledge. The ability to collect, understand and integrate such measurements at the conceptual level is fundamental to personalised or “precision” medicine. To aid researchers to connect these different pieces of information and reason over it, here we present our work on modeling of case report forms for COVID-19, then on an ontology for representing quantitative traits, and finally on integration within PhenoPackets, an exchange standard for the transmission of patient phenotype data, to showcase its applicability.

## Results

### Mapping to ontologies

We used the WHO COVID-19 case report form (CRF) RAPID version 23 March 2020^1^ to manually extract quantitative traits relevant for COVID-19. This CRF is divided in three modules: 1) admission to healthcare; 2) admission to ICU and follow-up patients; and 3) outcome. We extracted quantitative traits and units terms from modules 1 and 2 (located in the *vital signs* and *laboratory results* sections). Following recommendations by domain-experts immunologists with experience of COVID-19 patients, the initial list was augmented with further traits, reaching a final total of 57 trait terms. The CMO, an OBO Foundry ontology designed to define phenotype measurement data, has the precise concepts to annotate 67% of terms. We annotated the rest of terms using the Experimental Factor Ontology (EFO) [2] and the NCI Thesaurus (NCIT) [3]. Units were annotated using the Units Ontology (UO) [4], where 63% of terms are represented. We then extracted and annotated patient metadata to capture mainly comorbidities, pre-admission medication and treatment terms, which were prioritized as clinical metadata for future application on sequence submission. Comorbidities were annotated with the Disease Ontology (79%) [5], the Human Phenotype Ontology [6] and EFO, and medications using mainly ChEBI (65%) [7] and NCIT.

### Lexical mining of CMO labels and axiom patterns

We lexically decomposed the labels of CMO classes using the labels of the Uberon [8], ChEBI, and PATO [9] ontologies, following a strategy similar to the one previously applied to extract mappings from Gene Ontology labels [10]^2^. We built a dictionary from the labels and synonyms of classes in Uberon, ChEBI, and PATO, and identified their occurrence in the labels of CMO classes. For example, the CMO class “heart measurement” (CMO:0000670) matches with the Uberon class “heart” (UBERON:0000948).

We then added OWL axioms to the CMO based on the patterns used to define phenotype ontologies [11] and integrated the CMO with the Mammalian Phenotype Ontology (MP) [12]. Using the ELK reasoner [13] we were then able to infer relations between measurements and the phenotypes that may be detected using these measurements. For example, the CMO class “heart measurement” may be used to detect phenotypes falling under the MP classes “abnormal heart morphology” (MP:0000266) and “abnormal cardiovascular system physiology” (MP:0001544).

Source code is freely available at https://github.com/leechuck/qto.

### Quantitative trait data model

From the list of semantically annotated quantitative traits and units, we developed a data model which enables quantitative trait data to be machine-readable and interoperable for sharing and amenable to bioinformatics analysis. The data model is based on the Information Artifact Ontology (IAO) [14], the BioAssay Ontology (BAO) [15], the Ontology for Biomedical Investigations (OBI) [16] and UO. We use CMO classes to define the types and we implemented the Shape Expressions (ShEx) shape^3^ to communicate the graph structure, generate and validate the instance data. Finally, we would like to highlight the knowledge gap detected in the biomedical ontological space for COVID-19 related clinical concepts thus offering an opportunity to expand and enrich viral-related content for CMO and UO. For example the cytokine measurement “IL-8 pg/mL”, we couldn’t find a term neither in CMO nor in UO. Data model and ShEx shape are freely available at https://github.com/NuriaQueralt/BioHackathon/tree/master/bh20-ontology-qt.

### Integration into GA4GH PhenoPackets standard

PhenoPackets is an exchange standard for packaging observable indicators and phenotype data together into an interlinked model of disease, patient data and corresponding characteristic values of traits. Phenotypes, indicators and disease expression are not symptomatically linked, but mediated by patients (in biomedical context) or organisms acting as key links. PhenoPackets model the interlinking and enable quantitative analysis of these interactions. Based on the GA4GH inlined PhenoPackets schema^4^, an extension to integrate the quantitative trait data model with additional trait ontologies and data was defined. This schema provides a “QuantitativeTraitFeature” base element to characterize and attach trait features and observable data. Conforming with the PhenoPackets reference implementation, Protobuf-based serialization was implemented to facilitate interoperable exchange with biomedical databases, repositories as well as computational pipelines. It also enables integration of phenotypic and quantitative data with sequence data. Future work will aim at applying this quantitative trait model to many other use-cases and applications.

Source code for the PhenoPackets extension is freely available at https://github.com/cp-weiland/QuantitativeTraitFeature.

## Data Availability

Source code is freely available at https://github.com/leechuck/qto
Data model and ShEx shape are freely available at
https://github.com/NuriaQueralt/BioHackathon/tree/master/bh20-ontology-qt
Source code for the PhenoPackets extension is freely available at
https://github.com/cp-weiland/QuantitativeTraitFeature

https://github.com/leechuck/qto

https://github.com/NuriaQueralt/BioHackathon/tree/master/bh20-ontology-qt/tree/master/bh20-ontology-qt

https://github.com/cp-weiland/QuantitativeTraitFeature

## Footnotes

1 https://www.who.int/docs/default-source/coronaviruse/who-ncov-crf.pdf?sfvrsn=84766e69_2

2 https://protege.stanford.edu/conference/2006/submissions/demos/bada_demo%20description.pdf

3 https://github.com/shexSpec/shex/wiki/ShEx

4 https://github.com/phenopackets/phenopacket-schema

